# Regional Disparities in the Social Determinants of Stroke Severity

**DOI:** 10.1101/2025.05.06.25327114

**Authors:** Jacobus Barnard, David Lin, Grace Armstrong, Neon Calumpang, Miranda Cash, Amy Han

## Abstract

**Introduction:** Social determinants of health (SDOH) are known factors of stroke risk and outcomes. We aim to gain a comprehensive understanding of the impact of stroke risk-specific SDOH on stroke severity by investigating the patient population served by a regional healthcare system.

**Methods:** This retrospective cohort study analyzed patient data from the American Heart Association’s Get With The Guidelines-Stroke (AHA GWTG-S) Case Record Forms, collected from three stroke centers between January 2022 and May 2024. We compared the patient ordinal modified Rankin Scale (mRS) and the National Institutes of Health Stroke Scale (NIHSS) scores to the predictor variables of age, sex, race, Hispanic ethnicity, ZIP code, payment sources, and mode of arrival.

**Results:** When age-adjusted, Black or African American patients had higher mean NIHSS scores and higher rates of Minor Stroke Symptoms than White patients (*p*<0.01, *p*<0.01). ZIP Codes with higher mean NIHSS stroke scale scores correlated significantly with ZIP Codes defined by lower median household income (r=-0.61, *p*<0.01), lower education attainment (r=-0.71, *p*<0.001), and higher percentages of minority group populations (r=0.50, *p*<0.02). Patients with higher mean scores across all measures were those associated with using Medicare versus private insurance (*p*<0.0001) and those arriving at the hospital via EMS versus private transport (*p*<0.01).

**Conclusions:** This research addresses the significance of surveying region-specific social determinants of health for insight into targeted interventions. Initiatives such as expanding stroke awareness education and increasing preventative screenings in the community may reduce disparities in stroke severity and improve outcomes in underserved areas.

## INTRODUCTION

### I. Background

Strokes, or cerebrovascular accidents, are severe medical emergencies responsible for 1 of every 21 U.S. deaths—approximately one every 3 minutes and 17 seconds.^1^ Cerebrovascular diseases are the 5^th^ leading cause of death in the U.S.;^2^ to a greater extent, they are the 3^rd^ leading cause of death and disability combined worldwide.^3^ Stroke incidents have been estimated to occur in 9.4 million U.S. adults over 20 years of age, a prevalence of 3.3%, based on 2017–March 2020 Pre-Pandemic self-reported NHANES data.^4,5^ Despite the national average of stroke mortality having declined from 1968–2016 with the improvement of medical care, geographical disparities exist across states and their counties such as Lake County, Indiana.^1,4,5^ From 2019–2021, Lake County averaged 79 stroke deaths per 100,000, rising to 123 per 100,000 among Black or African American patients—among the highest rates statewide.^6^ These averages are markedly increased compared to the national average of 41.4 deaths per 100,000.^2^ These disparities highlight a critical need to investigate the underlying determinants of health contributing to this inequity.

Stroke risk is attributed to modifiable risk factors such as hypertension, obesity, hyperglycemia, and hyperlipidemia; co-morbidities such as coronary heart disease, renal disease, and liver disease; and behavioral risk factors such as smoking, physical activity, and nutrition.^1^ As of 2021, the Lake County, had high rates of hypertension (37.8%), obesity (40.5%), high cholesterol (36.8%), diagnosed diabetes (11.0%), smoking (18.2%), and physical inactivity (23.3%).^7^ The prevalence of obesity and smoking rates in Lake County are higher than the state averages, being 36.3% and 17.3%, respectively, which could be contributing to the county’s increased stroke incidence and mortality.^8,9^

Social determinants of health (SDOH) are underlying factors that strongly influence individual health-related stroke risk factors and outcomes. The World Health Organization (WHO) has defined SDOH as 1) the conditions of daily life, encompassed by the circumstances in which people are born, grow up, live, work, and age; 2) the structural drivers of those conditions (as shaped by the distribution of power, prevalence of discrimination, and access to resources); and 3), the systems established and available to support health and deal with illness.^10^ Social determinants of risk for cardiovascular disease are an interrelated group of factors regarding socioeconomic position (describing education and income), race and ethnicity, access to care, social support, residential environment, as well as culture and language.^11^ Prior studies have shown that lower socioeconomic status (SES) results in higher stroke incidence in elderly patients (≥65 years), higher odds of post-stroke disability for all ages, increased hospital length of stay for minority groups, and higher risk of stroke and myocardial infarction mortality.^12^ In one 2022 nationwide geological analysis of county-level SDOH, it was found that higher stroke hospitalization rates were associated with heart disease prevalence, blood pressure medication nonadherence, age-adjusted obesity, hospital access to neurological services, and female head of household.^12^ Supplemental Figure S1 illustrates heatmaps of Lake County, IN generated from U.S. Census Bureau data, available in Supplemental Table 1, to contextualize the non-homogenous geographical distribution of wealth, education, and of minority race and ethnic group populations.^13,14^

### II. Objectives

Stroke Centers and related stroke healthcare units are facilities certified to deliver varying degrees of stroke care. Powers Health (formerly Community Healthcare Systems) is a reputable Lake County, IN healthcare system and hospital network. They deliver stroke care at their Joint Commission Certified Comprehensive Stroke Center (CSC) at Community Hospital in Munster and are supported by the certified Primary Stroke Centers (PSCs) at St. Catherine Hospital in East Chicago and St. Mary Medical Center in Hobart. Depending on the complexity of the acute stroke presentation and the resources available, PSCs and acute stroke-ready hospitals often coordinate patient hospital transfers to higher-level stroke centers, such as CSCs. The 2025 Joint Commission organization listings state that there are only three accredited CSCs in Indiana.^15^ Community Hospital, under Powers Health, is one CSC that provides service to Lake County, which has the highest state-level stroke incidence, and serves as an ideal case study and community partner to analyze a regional patient population for stroke disparities.

The Joint Commission mandates the evaluation and documentation of admitted stroke patients using standardized clinical stroke scales. The modified Rankin Scale (mRS) is a widely used clinician-reported measure with evidenced value as an end-point for randomized clinical stroke control trials and is a useful instrument in assessing recovery from stroke.^16^ mRS evaluates the physical capabilities of a patient on a scale of 0-6, with a higher score correlating with more functional deficits. The National Institute of Health Stroke Scale (NIHSS) is a detailed clinical measurement of stroke symptom severity that assesses neurological function. NIHSS scores range from 0-42, with a higher score correlating with more neurological deficits.^17^ While the mRS is used for the assessment of stroke recovery and function, the baseline NIHSS is a strong predictor of outcomes after stroke.^18^ Notably, disparities seen in baseline NIHSS scores can be attributed to prehospital factors, such as stroke symptom recognition and emergency response time.^19^ Both the mRS and NIHSS provide valuable insight as clinical measurements in assessing the stroke severity of the patients in this study. Score categories for both clinical scales are available in Supplemental Table S2.

Northwest Indiana and its surrounding urban and rural areas make up a diverse patient population with high stroke rates. While generalizable associations of stroke risk-specific social determinants of health and stroke incidence have been shown in previous county-level studies, ZIP Code-level disparities in stroke severity are not well-characterized within regional stroke systems of care. Our study aims to investigate the specific patient population served by an individual hospital system by assessing the impact of stroke risk-specific social determinants on stroke incidence, severity, and outcomes.

## METHODS

### I. Study Design

This retrospective cohort study analyzed patient data from the American Heart Association’s Get With The Guidelines-Stroke (AHA GWTG-S) Case Record Forms.^20^ This study was approved by the Institutional Review Board at Indiana University School of Medicine (IRB 23783) and followed STROBE (Strengthening the Reporting of Observational Studies in Epidemiology) guidelines for observational research, available in Supplemental Materials.^21^

### II. Population

Between January 2022 and May 2024, a total of 2861 stroke patients from over 130 ZIP Codes across 22 counties were admitted to Community Hospital, St. Mary’s Hospital, and St. Catherine’s Hospital. To better represent the local population, ZIP Codes with fewer than five patients were excluded, limiting the sample size to within 50 miles of the CSC. This yielded a total for analysis to 2569 stroke patients from 46 ZIP Codes across 7 counties. Patients were further sub-set based on their Mode of Arrival to the hospital—either “Transfer from another hospital” or “Non-Transfer”— to minimize the statistical bias introduced by transfer patients having more severe, higher stroke scale scores upon intake. Two ecological ZIP-Code level analyses were performed:

1. The Focused (25 ZIP Code) Analysis: Included all 25 ZIP Codes of one county, Lake County, IN (n=1,563 Non-Transfer patients).
2. The Expanded (46 ZIP Code) Analysis: Included 46 ZIP Codes across seven counties, made up of Lake County, IN; Porter County, IN; Jasper County, IN; LaPorte County, IN; Starke County, IN; Will County, IL; and Cook County, IL. This analysis comprises a total of 2087 Non-Transfer patients from 41 ZIP Codes and 594 Transfer patients from 43 ZIP Codes.

### II. Data Elements

Data abstraction was conducted by the Power Health’s Neuroscience Research department and the CSC staff. Inclusion was based on patients’ Principal Diagnosis ICD-10-CM codes (I62-I69, G45). Patients with documented stroke scores were also included independent of their Principal Diagnosis ICD-10-CM code. One author had full access to all the data in the study and takes responsibility for its integrity and the data analysis.

The predictor variables include data elements available from the AHA GWTG-S Case Record Forms. Predictor variables included patient Age (categorized into one of eight age groups; <18, 18-24, 25-39, 40-49, 50-64, 65-74, 75-84, >84), self-reported Race and Ethnicity (non-Hispanic Black or African American, non-Hispanic White, Hispanic, Asian, and non-Hispanic American Indiana/ Alaska Native), ZIP Code, Insurance Type, and Mode of Arrival (EMS or private transport). Patients who identified as Hispanic ethnicity were grouped into the Hispanic Ethnicity category, regardless of their other racial identities. ZIP Code-level demographic variables such as Median Household Income, Education Level, and Minority Population were derived from U.S. Census Bureau ZCTA5 statistics as ecological proxies since individual-level income and education data were not available.

The outcome measures of interest were the reported ordinal scores of the modified Rankin Scale (mRS) and the National Institute of Health Stroke Scale (NIHSS), with scores taken at two points in the patient’s timeline of care. Assessing mRS and NIHSS values at different time points along a stroke patient’s timeline of care provides information about an intervention’s efficacy.^22^ In our study, the two mRS data points are captured as “Pre-Stroke mRS and “Discharge mRS,” describing the patient’s functional ability before stroke onset and at hospital discharge. Our two NIHSS data points are captured as “Initial NIHSS” and “Discharge NIHSS,” describing the patient’s neurological ability shortly after stroke onset and at hospital discharge.

### III. Instrumentation, Analysis, and Checks (Ensuring Validity and Reliability)

Descriptive statistics (mean, median ± inter-quartile range (IQR)) were used for ordinal (nonparametric) data. Percentages were calculated for incidence rates. Results were visualized using box plots, bar graphs, scatter plots, and heatmaps. ZIP Code shapefile data used to create the heatmap figures was obtained from the ArcGIS Hub database.^23^ Variables were sorted by ordinal counts and age category (<18, 18-24, 25-39, 40-49, 50-64, 65-74, 75-84, >84); select variables underwent age-adjustment via a direct standardization technique using the World (WHO 2000-2025) Standard Population proportions.^24^ Patients with incomplete NIHSS or mRS scores were excluded.

The Mann-Whitney U (Wilcoxon) rank-based test assessed the ordinal differences between two predictor variable groups. The Kruskal-Wallis test was performed for comparisons between more than two groups. Pending any statistical significance, a follow-up Dunn post hoc test was performed for pairwise comparisons using a Bonferroni correction method to control for the increased likelihood of Type I errors. Chi-squared analysis was conducted on incidence data; this specifically included variables of Sex and Race and Ethnicity. Pearson correlation coefficients were also obtained to quantify the strength and direction of a linear relationship.

All statistical analyses were performed using R (version 4.1.1).^25^

## FOCUSED (25 ZIP CODE) ANALYSIS RESULTS

Descriptive statistics and bivariate analysis results are reported in Table 1.

**Table 1.**
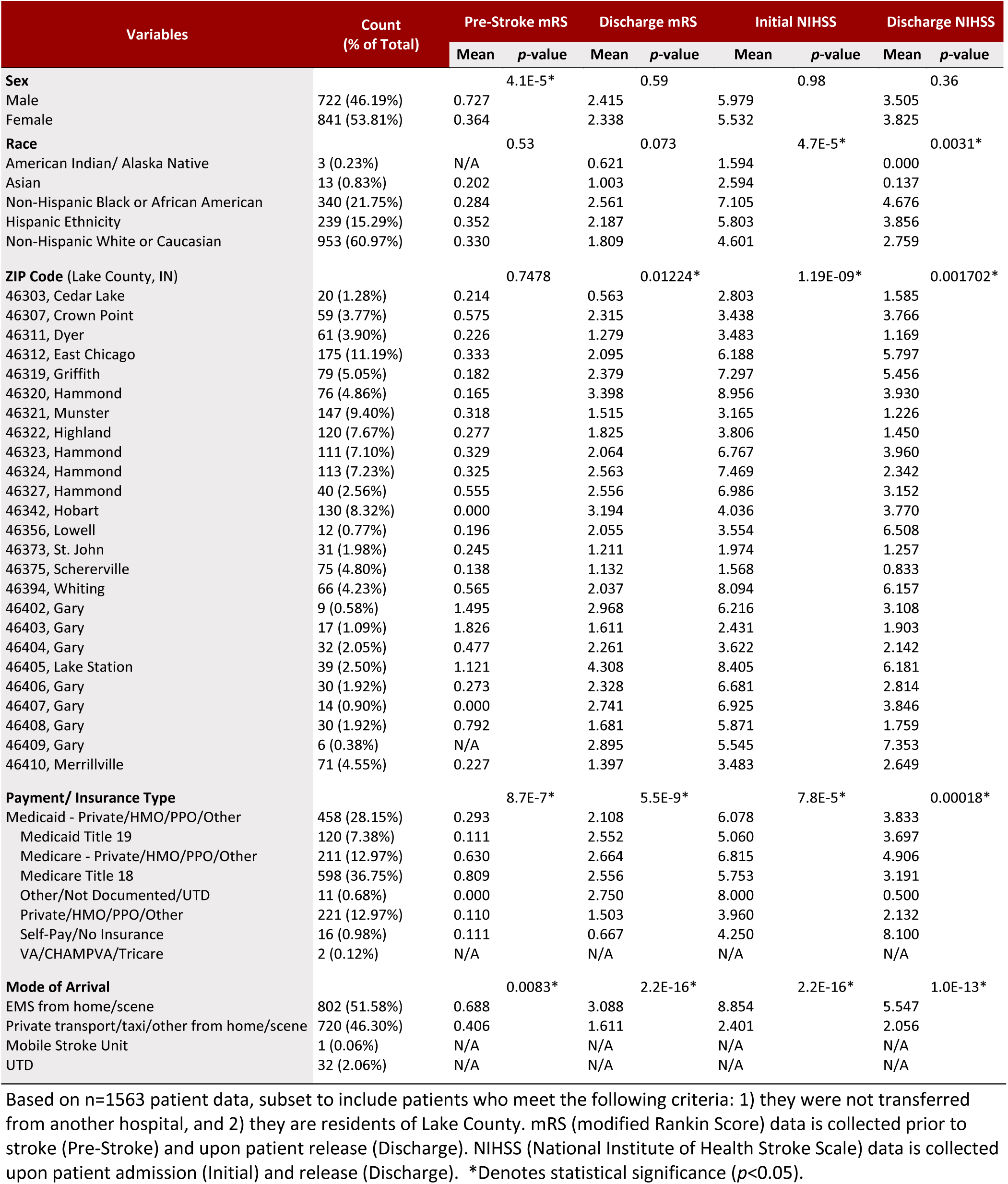
Descriptive Statistics of Patient Population and Bivariate Analysis of mRS and NIHHS Scores by Select Social Determinant of Health Variables.

| Variables | Count<br>(% of Total) | Pre-Stroke mRS |  | Discharge mRS |  | Initial NIHSS |  | Discharge NIHSS |  |
| --- | --- | --- | --- | --- | --- | --- | --- | --- | --- |
|  |  | Mean | p-value | Mean | p-value | Mean | p-value | Mean | p-value |
| **Sex** |  |  | 4.1E-5* |  | 0.59 |  | 0.98 |  | 0.36 |
| Male | 722 (46.19%) | 0.727 |  | 2.415 |  | 5.979 |  | 3.505 |  |
| Female | 841 (53.81%) | 0.364 |  | 2.338 |  | 5.532 |  | 3.825 |  |
| **Race** |  |  | 0.53 |  | 0.073 |  | 4.7E-5* |  | 0.0031* |
| American Indian/ Alaska Native | 3 (0.23%) | N/A |  | 0.621 |  | 1.594 |  | 0.000 |  |
| Asian | 13 (0.83%) | 0.202 |  | 1.003 |  | 2.594 |  | 0.137 |  |
| Non-Hispanic Black or African American | 340 (21.75%) | 0.284 |  | 2.561 |  | 7.105 |  | 4.676 |  |
| Hispanic Ethnicity | 239 (15.29%) | 0.352 |  | 2.187 |  | 5.803 |  | 3.856 |  |
| Non-Hispanic White or Caucasian | 953 (60.97%) | 0.330 |  | 1.809 |  | 4.601 |  | 2.759 |  |
| **ZIP Code (Lake County, IN)** |  |  | 0.7478 |  | 0.01224* |  | 1.19E-09* |  | 0.001702* |
| 46303, Cedar Lake | 20 (1.28%) | 0.214 |  | 0.563 |  | 2.803 |  | 1.585 |  |
| 46307, Crown Point | 59 (3.77%) | 0.575 |  | 2.315 |  | 3.438 |  | 3.766 |  |
| 46311, Dyer | 61 (3.90%) | 0.226 |  | 1.279 |  | 3.483 |  | 1.169 |  |
| 46312, East Chicago | 175 (11.19%) | 0.333 |  | 2.095 |  | 6.188 |  | 5.797 |  |
| 46319, Griffith | 79 (5.05%) | 0.182 |  | 2.379 |  | 7.297 |  | 5.456 |  |
| 46320, Hammond | 76 (4.86%) | 0.165 |  | 3.398 |  | 8.956 |  | 3.930 |  |
| 46321, Munster | 147 (9.40%) | 0.318 |  | 1.515 |  | 3.165 |  | 1.226 |  |
| 46322, Highland | 120 (7.67%) | 0.277 |  | 1.825 |  | 3.806 |  | 1.450 |  |
| 46323, Hammond | 111 (7.10%) | 0.329 |  | 2.064 |  | 6.767 |  | 3.960 |  |
| 46324, Hammond | 113 (7.23%) | 0.325 |  | 2.563 |  | 7.469 |  | 2.342 |  |
| 46327, Hammond | 40 (2.56%) | 0.555 |  | 2.556 |  | 6.986 |  | 3.152 |  |
| 46342, Hobart | 130 (8.32%) | 0.000 |  | 3.194 |  | 4.036 |  | 3.770 |  |
| 46356, Lowell | 12 (0.77%) | 0.196 |  | 2.055 |  | 3.554 |  | 6.508 |  |
| 46373, St. John | 31 (1.98%) | 0.245 |  | 1.211 |  | 1.974 |  | 1.257 |  |
| 46375, Schererville | 75 (4.80%) | 0.138 |  | 1.132 |  | 1.568 |  | 0.833 |  |
| 46394, Whiting | 66 (4.23%) | 0.565 |  | 2.037 |  | 8.094 |  | 6.157 |  |
| 46402, Gary | 9 (0.58%) | 1.495 |  | 2.968 |  | 6.216 |  | 3.108 |  |
| 46403, Gary | 17 (1.09%) | 1.826 |  | 1.611 |  | 2.431 |  | 1.903 |  |
| 46404, Gary | 32 (2.05%) | 0.477 |  | 2.261 |  | 3.622 |  | 2.142 |  |
| 46405, Lake Station | 39 (2.50%) | 1.121 |  | 4.308 |  | 8.405 |  | 6.181 |  |
| 46406, Gary | 30 (1.92%) | 0.273 |  | 2.328 |  | 6.681 |  | 2.814 |  |
| 46407, Gary | 14 (0.90%) | 0.000 |  | 2.741 |  | 6.925 |  | 3.846 |  |
| 46408, Gary | 30 (1.92%) | 0.792 |  | 1.681 |  | 5.871 |  | 1.759 |  |
| 46409, Gary | 6 (0.38%) | N/A |  | 2.895 |  | 5.545 |  | 7.353 |  |
| 46410, Merrillville | 71 (4.55%) | 0.227 |  | 1.397 |  | 3.483 |  | 2.649 |  |
| **Payment/ Insurance Type** |  |  | 8.7E-7* |  | 5.5E-9* |  | 7.8E-5* |  | 0.00018* |
| Medicaid - Private/HMO/PPO/Other | 458 (28.15%) | 0.293 |  | 2.108 |  | 6.078 |  | 3.833 |  |
| Medicaid Title 19 | 120 (7.38%) | 0.111 |  | 2.552 |  | 5.060 |  | 3.697 |  |
| Medicare - Private/HMO/PPO/Other | 211 (12.97%) | 0.630 |  | 2.664 |  | 6.815 |  | 4.906 |  |
| Medicare Title 18 | 598 (36.75%) | 0.809 |  | 2.556 |  | 5.753 |  | 3.191 |  |
| Other/Not Documented/UTD | 11 (0.68%) | 0.000 |  | 2.750 |  | 8.000 |  | 0.500 |  |
| Private/HMO/PPO/Other | 221 (12.97%) | 0.110 |  | 1.503 |  | 3.960 |  | 2.132 |  |
| Self-Pay/No Insurance | 16 (0.98%) | 0.111 |  | 0.667 |  | 4.250 |  | 8.100 |  |
| VA/CHAMPVA/Tricare | 2 (0.12%) | N/A |  | N/A |  | N/A |  | N/A |  |
| **Mode of Arrival** |  |  | 0.0083* |  | 2.2E-16* |  | 2.2E-16* |  | 1.0E-13* |
| EMS from home/scene | 802 (51.58%) | 0.688 |  | 3.088 |  | 8.854 |  | 5.547 |  |
| Private transport/taxi/other from home/scene | 720 (46.30%) | 0.406 |  | 1.611 |  | 2.401 |  | 2.056 |  |
| Mobile Stroke Unit | 1 (0.06%) | N/A |  | N/A |  | N/A |  | N/A |  |
| UTD | 32 (2.06%) | N/A |  | N/A |  | N/A |  | N/A |  |

Based on n=1563 patient data, subset to include patients who meet the following criteria: 1) they were not transferred from another hospital, and 2) they are residents of Lake County. mRS (modified Rankin Score) data is collected prior to stroke (Pre-Stroke) and upon patient release (Discharge). NIHSS (National Institute of Health Stroke Scale) data is collected upon patient admission (Initial) and release (Discharge). \*Denotes statistical significance ( $p < 0.05$ ).

### I. Sex

Non-age-adjusted Mann-Whitney U tests per stroke scale, available in Supplemental Table S3, reveal no significant differences between sexes for Initial NIHSS (*p*=0.98), Discharge mRS (*p*=0.59), and Discharge NIHSS scores (*p*=0.36). However male patients, with a mean Pre-Stroke mRS score of 0.7237, had a 0.36 higher mean score than female patients (*p*=0.000041), as illustrated in Supplemental Figure S2.

Age-adjusted Chi-square tests of Initial NIHSS scores organized into categories of severity reveal that male patients had a 14.85% higher incidence rate of Moderate Stroke severity than female patients (*p*=0.0102). Descriptive statistics are detailed in Supplemental Table S4, and follow-up pairwise comparisons are shown in Supplemental Table S5.

Age-adjusted Chi-square tests of Discharge NIHSS scores organized into categories of severity reveal no significant differences in incidence rates between categories. Descriptive statistics are detailed in Supplemental Table S6, and follow-up pairwise comparisons are shown in Supplemental Table S7.

### II. Race and Ethnicity

Age-adjusted Kruskal Wallis tests per stroke scale, available in Supplemental Table S8, reveal no significant differences between Race and Ethnicity groups for Pre-Stroke mRS (*p*=0.53) and Discharge mRS (*p*=0.07), but does display significant differences in Initial NIHSS scores (*p*=0.0047) and Discharge NIHSS scores (*p*=0.0031). A follow-up Dunn test of Initial NIHSS scores, available in Supplemental Table S9, reveals that Black or African American patients, with a mean Initial NIHSS score of 7.1049, have a 2.5-point higher mean score than White patients (*p*=0.000098). A follow-up Dunn test of Discharge NIHSS scores, available in Supplemental Table S10, reveals that Black or African American patients, with a mean Discharge NIHSS score of 4.6755, have a 1.916 point higher mean score than White patients (*p*=0.00397). No other cross-comparisons had observed significant differences between Race and Ethnicity groups. The spread of Initial and Discharge NIHSS scores between Race and Ethnicity groups is available in Supplemental Figure S3.

Age-adjusted Chi-square tests of Initial NIHSS scores organized into categories of severity reveal non-specific significant differences between Race and Ethnicity groups in their incidence rates of No Stroke Symptoms severity (*p*=0.0065), Minor Stroke severity (*p*=0.0487), and Moderate Stroke severity (*p=*0.0135). When carrying out specific pairwise comparisons: White patients had an incidence rate of No Stroke Symptoms 14.17% higher than Hispanic Ethnicity patients (*p* =0.0194) and 13.85% higher than Black or African American patients (*p*=0.0055); Black or African American patients had an incidence rate of Minor Stroke severity 31.81% higher than Hispanic Ethnicity patients (*p*=0.0013); White patients had an incidence rate of Moderate Stroke severity 12.12% higher than Hispanic Ethnicity patients (*p*=0.0405). Descriptive statistics are detailed in Supplemental Table S10, and follow-up pairwise comparisons are shown in Supplemental Table S11. No other cross-comparisons had observed significant differences between Race and Ethnicity groups. Figure 1A displays the incidence of Initial NIHSS stroke scores organized into categories of severity by Race and Ethnicity groups.

**Figure 1.**
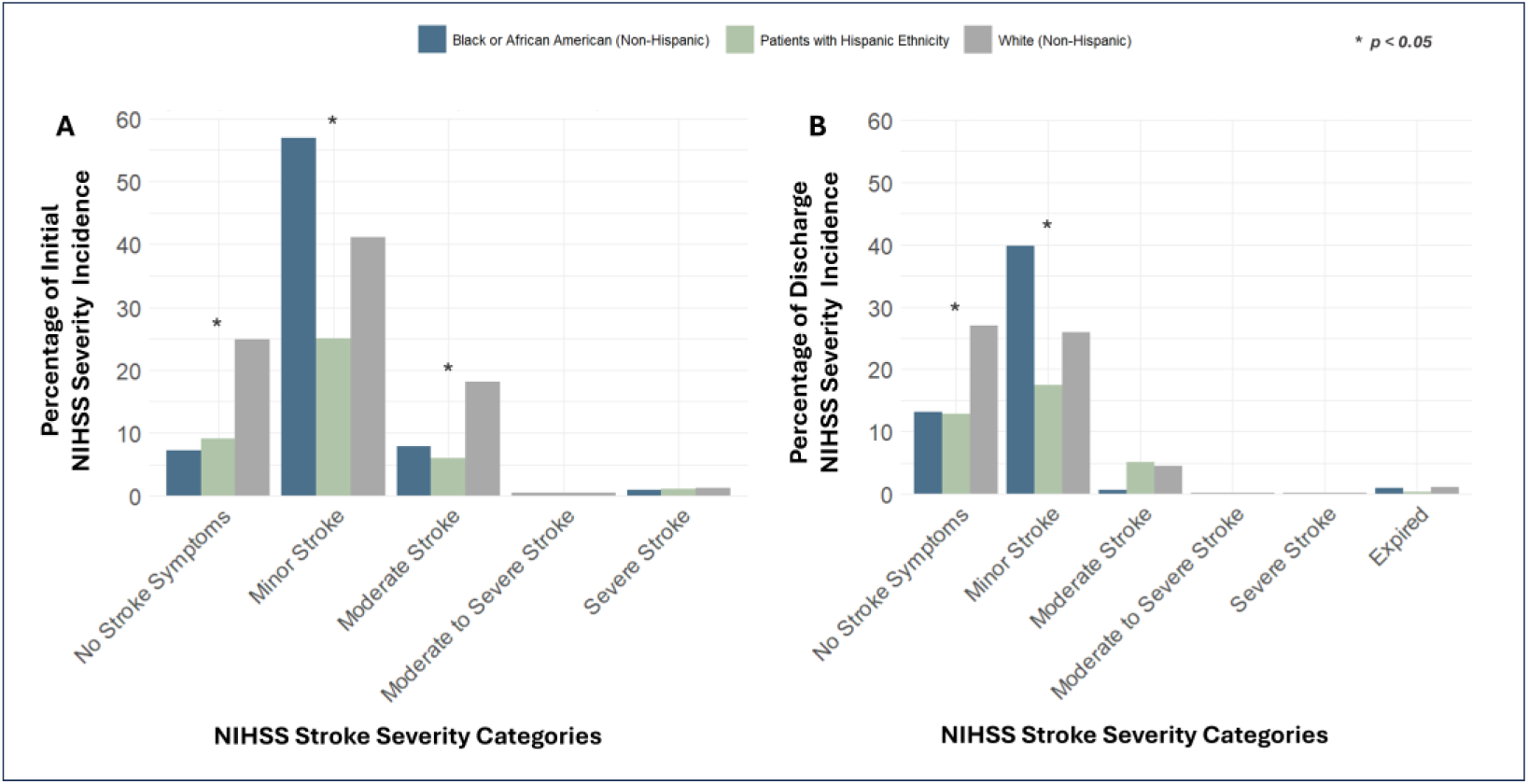
Distribution of NIHSS Stroke Severity Categories by Race and Ethnicity, Age-Adjusted. Illustrates the percentage incidence of patients, grouped by Race and Ethnicity and adjusted for age, in each NIHSS (National Institute of Health Stroke Scale) stroke severity category upon patient admission (Initial) and release (Discharge). Differences in proportions between groups were analyzed using the Chi-squared test. *Denotes statistical signiiccance (*p*<0.05).

Age-adjusted Chi-square tests of Discharge NIHSS scores organized into categories of severity reveal non-specific significant differences between Race and Ethnicity groups in their incidence rates of Minor Stroke severity (*p*=0.0106). When carrying out specific pairwise comparisons, Black or African American patients had an incidence rate of Moderate Stroke severity 31.81% higher than Hispanic Ethnicity patients (*p*=0.0098); meanwhile, White patients had an incidence rate of Minor Stroke severity 22.26% higher than Hispanic Ethnicity patients (*p*=0.0405). Descriptive statistics are detailed in Supplemental Table S11, and follow-up pairwise comparisons are shown in Supplemental Table S13. No other cross-comparisons had observed significant differences between Race and Ethnicity groups. Figure 1B displays the incidence of Discharge NIHSS stroke scores organized into categories of severity by Race and Ethnicity groups.

### III. ZIP Code

Table 2 details the results of age-adjusted Kruskal Wallis tests per stroke scale and any significant follow-up pairwise comparison results. Full descriptive statistics are detailed in Supplemental Table S14, and all follow-up pairwise comparisons are shown in Supplemental Table S15. Heatmaps displaying ZIP Code-level results are illustrated in Supplemental Figure S4.

**Table 2.** Focused (25 ZIP Code) Descriptive, Kruskal Wallis, and Significant Pairwise Comparison Statistics for mRS and NIHSS by ZIP Code, Age-Adjusted.

| Clinical Stroke Measure Variable |  |  |  |  |  |  |  |  |  |
| --- | --- | --- | --- | --- | --- | --- | --- | --- | --- |
|  | Kruskal Wallis Test |  |  | Significant Pairwise Comparisons |  |  |  |  |  |
|  | KW X2 Statistic | Degrees of Freedom | p-value | ZIP Codes | Municipality | n | Mean | Difference | p-value |
| Pre-Stroke mRS |  |  |  |  |  |  |  |  |  |
|  | 18.178 | 23 | 0.7478 | N/A |  |  |  |  |  |
| Discharge mRS |  |  |  |  |  |  |  |  |  |
|  | 42.2050 | 24 | 0.01224 | 46320 (Ref.) | Hammond | 69 | 3.398 |  |  |
|  |  |  |  | 46303 | Cedar Lake | 17 | 0.563 | - 2.835 | 0.0464 |
|  |  |  |  | 46375 | Schererville | 72 | 1.132 | - 2.266 | 0.0451 |
| Initial NIHSS |  |  |  |  |  |  |  |  |  |
|  | 89.2645 | 24 | 1.19E-09 | 46375 (Ref.) | Schererville | 70 | 1.568 |  |  |
|  |  |  |  | 46320 | Hammond | 73 | 8.956 | + 7.388 | 0.031 |
|  |  |  |  | 46394 | Whiting | 66 | 8.094 | + 6.526 | 0.042 |
|  |  |  |  | 46319 | Griffith | 77 | 7.297 | + 5.729 | 6.1E-04 |
|  |  |  |  | 46323 | Hammond | 108 | 6.676 | + 5.199 | 4.1E-05 |
|  |  |  |  | 46312 | East Chicago | 163 | 6.188 | + 4.620 | 9.4E-05 |
|  |  |  |  | 46342 | Hobart | 123 | 4.036 | + 2.468 | 0.044 |
|  |  |  |  | 46321 (Ref.) | Munster | 139 | 3.165 |  |  |
|  |  |  |  | 46319 | Griffith | 77 | 7.297 | + 4.132 | 0.0167 |
|  |  |  |  | 46323 | Hammond | 108 | 6.676 | + 3.602 | 0.001 |
|  |  |  |  | 46312 | East Chicago | 163 | 6.188 | + 3.023 | 0.0022 |
|  |  |  |  | 46307 (Ref.) | Crown Point | 53 | 3.438 |  |  |
|  |  |  |  | 46323 | Hammond | 108 | 6.676 | + 3.329 | 0.0459 |
| Discharge NIHSS |  |  |  |  |  |  |  |  |  |
|  | 49.3690 | 24 | 0.001702 | 46312 (Ref.) | East Chicago | 83 | 5.797 |  |  |
|  |  |  |  | 46321 | Munster | 67 | 1.226 | - 4.571 | 0.014 |
|  |  |  |  | 46375 | Schererville | 43 | 0.833 | - 4.964 | 0.0016 |

Based on n=1563 patient data, subset to include patients who meet the following criteria: 1) they were not transferred from another hospital, and 2) they are residents of Lake County. Reference (Ref.) ZIP Codes were selected as such to have the least number of total reference groups. Differences among ZIP Code groups were assessed using the Kruskal-Wallis test. Post-hoc pairwise comparisons were performed using Dunn's test (with Bonferroni correction). mRS (modified Rankin Score) data is determined prior to stroke (Pre-Stroke) and upon patient release (Discharge). NIHSS (National Institute of Health Stroke Scale) data is collected upon patient admission (Initial) and release (Discharge).

Pearson’s correlation tests of stroke scales against select ZIP-code level SDOH statistics reveal significant correlations for Discharge mRS, Initial NIHSS, and Discharge NIHSS. Data is available in Supplemental Table S16. For Discharge mRS results, as stroke scores increase there was a moderate negative correlation between Median Household Income (*r*=-0.56, *p*=0.0038), a moderate negative correlation between Education Level (*r*=-0.60, *p*=0.00145), and a low positive correlation between Percent Minority Population (*r*=0.40, *p*=0.0467). For Initial NIHSS results, as stroke scores increase there was a moderate negative correlation between Median Household Income (*r*=-0.61, *p*=0.00119), a strong negative correlation between Education Level (*r*=-0.71, *p*=0.7853E-05), and a moderate positive correlation between Percent Minority Population (*r*=0.50, *p*=0.011). Correlation data for Initial NIHSS scores are demonstrated in Figure 2. For Discharge NIHSS results, as stroke scores increase, there was only found to be a low negative correlation with Education Level (*r*=-0.46, *p*=0.020).

**Figure 2.**
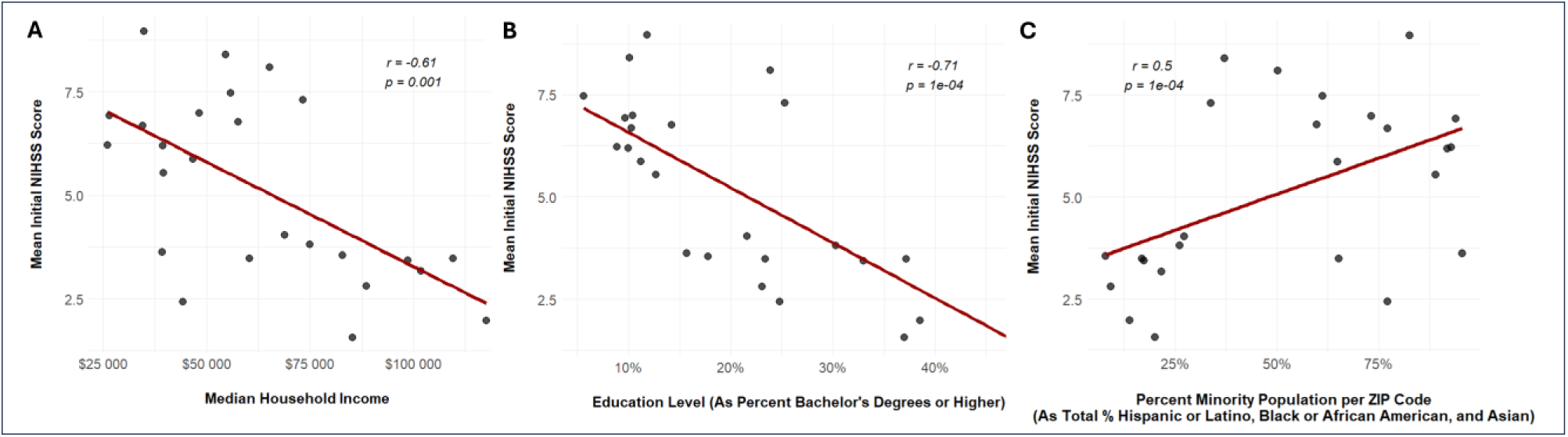
Scatter Plots of Select Social Determinants of Health vs. Mean Initial NIHSS Scores by ZIP Code, Age-Adjusted. Presents correlation between Initial NIHSS (National Institute of Health Stroke Scale) data, collected upon patient admission, and A) Median household income (in dollars), B) Education level (as percent bachelor’s degrees or higher), and C) Percent minority population. *r* is the correlation coefficient.

### IV. Payment Source

Age-adjusted Kruskal Wallis tests per stroke scale reveal significant differences in how patients paid and their Pre-Stroke mRS (*p*=8.05E-07), Initial NIHSS (*p*=7.76E-05), Discharge mRS (*p*=5.509E-09), and Discharge NIHSS scores (*p*=1.82E-04). Descriptive statistics are detailed in Supplemental Table S17, and follow-up pairwise comparisons are shown in Supplemental Table S18. Significant results are detailed in the following text.

Patients who paid with Medicare Title 18, with a mean Pre-Stroke mRS score of 0.809, have a mean Pre-Stroke mRS score 0.699 points higher than patients with Private/HMO/PPO/Other (*p*=0.0044) and 0.57 points lower than patients with Medicaid – Private/HMO/PPO/Other (*p*=2.55E-06). Patients with Private/HMO/PPO/Other, with a mean Pre-Stroke mRS Score of 0.110, have a mean Pre-Stroke mRS 0.52 points lower than patients with Medicare – Private/HMO/PPO/Other (*p*=0.00129).

Patients who paid with Medicare – Private/HMO/PPO/Other, with a mean Discharge mRS score of 2.664, have a mean Discharge mRS score 1.161 points higher than patients with Private/HMO/PPO/Other (*p*=9.06E-08) and 0.646 points higher than patients with Medicaid – Private/HMO/PPO/Other (*p*=0.027). Patients with Private/HMO/PPO/Other, with a mean Discharge mRS of 1.503, have a mean Discharge mRS score 1.049 points lower than patients with Medicaid Title 19 (*p*=0.0128), 1.053 points lower than patients with Medicare Title 18 (*p*=7.11E-07). Interestingly, patients who used Self-Pay/No Insurance, with a mean Discharge mRS score of 0.667, had a mean Discharge mRS score 2.553 points lower than those who paid with Medicare – Private/HMO/PPO/other (*p*=0.031).

Patients who paid with Private/HMO/PPO/Other, with a mean Initial NIHSS score of 3.960, have a mean Initial NIHSS score 2.118 points lower than patients with Medicaid – Private/HMO/PPO/Other (*p*=0.03) and 2.855 points lower than patients with Medicare – Private/HMO/PPO/Other (*p*=1.23E-05). Patients who paid with Medicare – Private/HMO/PPO/Other, with a mean Initial NIHSS score of 6.815, have a mean Initial NIHSS score 1.062 points higher than patients with Medicare Title 18 (*p*=0.0288).

Patients who paid with Medicare – Private/HMO/PPO/Other, with a mean Discharge NIHSS score of 4.606, have a mean Discharge NIHSS score 1.715 points higher than patients with Medicare Title 18 (*p*=0.0278) and 2.774 points higher than patients with Private/HMO/PPO/Other (*p*=3.65E-05).

### V. Mode of Arrival

Mann-Whitney U tests per stroke scale, available in Supplemental Table S19, reveal that patients coming from the home/scene who used EMS had significantly higher scores across all four clinical measures compared to patients who took Private transport/taxi/other. This result is further illustrated in Supplemental Figure S5. Patients who took EMS, with a mean Pre-Stroke mRS score of 0.688, have a Pre-Stroke mRS score 0.282 points higher than patients who took Private transport/taxi/other (*p*=0.0083). Patients who took EMS, with a mean Discharge mRS score of 3.088, have a Discharge mRS score 1.477 points higher than patients who took Private transport/taxi/other (*p=*2.20E-16). Patients who took EMS, with a mean Initial NIHSS score of 8.854, have a mean Initial NIHSS score 6.453 points higher than those who took Private transport/taxi/other (*p*=2.20E-16). Patients who took EMS, with a mean Discharge NIHSS score of 5.547, have a mean Initial NIHSS score 3.491 points higher than patients who took Private transport/taxi/other (*p*=1.0E-13).

## EXPANDED (46 ZIP CODE) ANALYSIS RESULTS

Descriptive statistics of patient transfer status are reported in Supplementary Table S20.

### I. ZIP Code (Non-Transfers)

Table 3 details the results of age-adjusted Kruskal Wallis tests per stroke scale and any significant follow-up pairwise comparison results. Full descriptive statistics are detailed in Supplemental Table S21, and all follow-up pairwise comparisons are shown in Supplemental Table S22. Heatmaps displaying ZIP Code-level results are illustrated in Figure 3.

**Figure 3.**
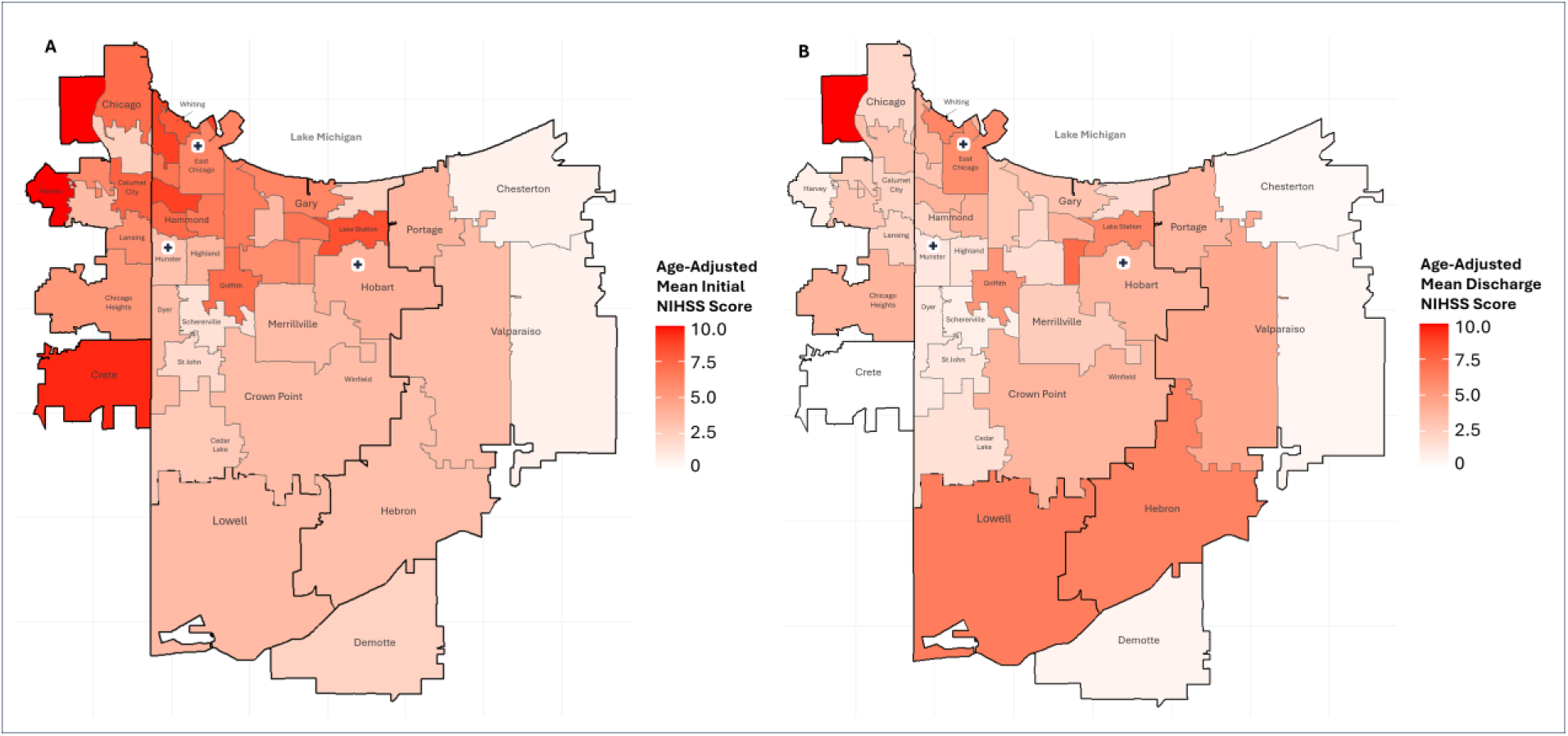
Heatmaps of Mean Stroke Scale Scores by ZIP Code, Age-Adjusted. A) Heatmap of age-adjusted mean Initial NIHSS (National Institute of Health Stroke Scale) scores; data collected upon patient admission. B) Heatmap of age-adjusted mean Discharge NIHSS scores; data collected upon patient release. Annotated with municipalities and markers indicating location of stroke centers.

**Table 3.**
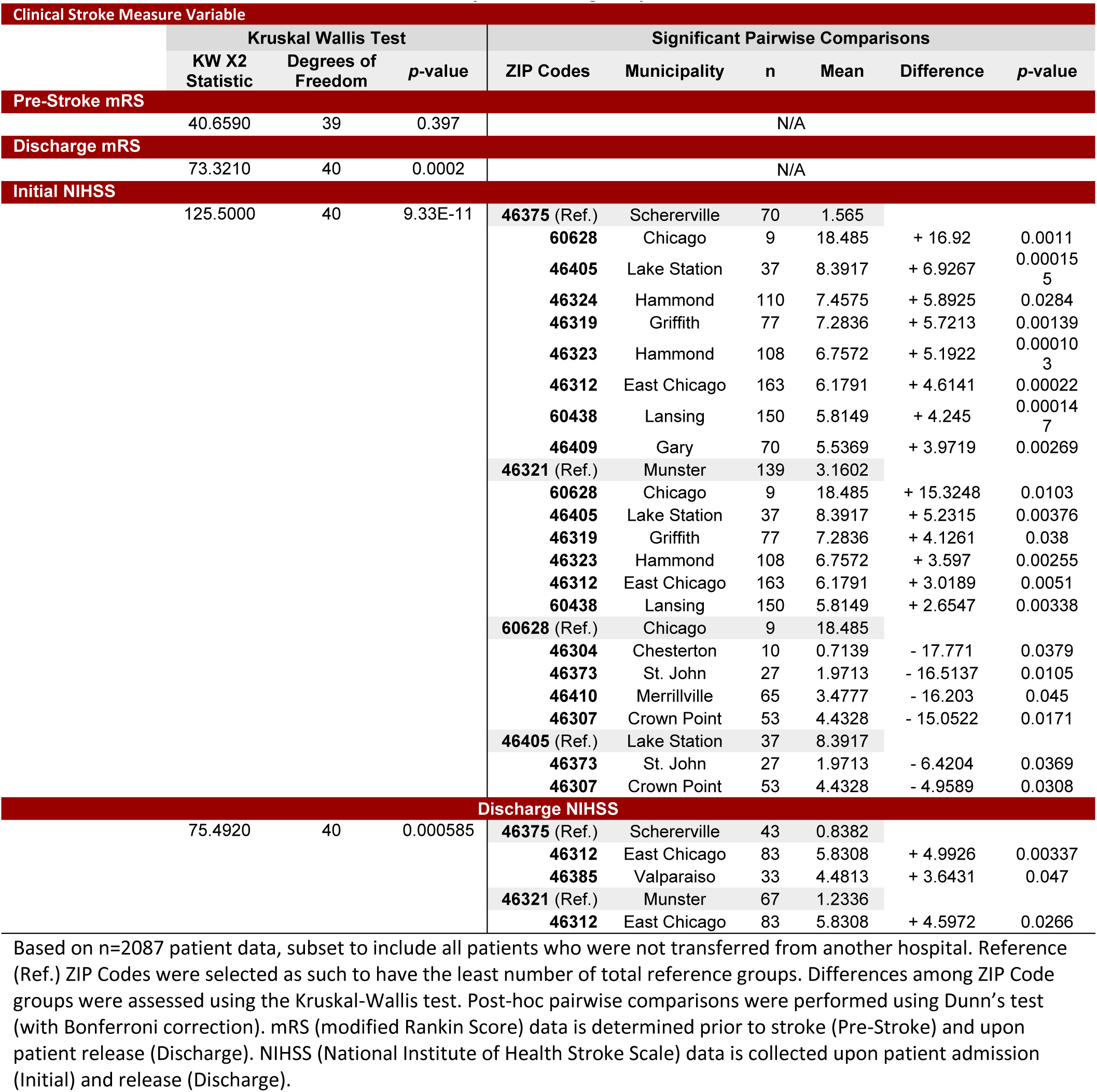
Expanded (46 ZIP Code) Descriptive, Kruskal Wallis, and Significant Pairwise Comparison Statistics for mRS and NIHSS by ZIP Code, Age-Adjusted.

| Clinical Stroke Measure Variable |  |  |  |  |  |  |  |  |  |
| --- | --- | --- | --- | --- | --- | --- | --- | --- | --- |
|  | Kruskal Wallis Test |  |  | Significant Pairwise Comparisons |  |  |  |  |  |
|  | KW X2 Statistic | Degrees of Freedom | p-value | ZIP Codes | Municipality | n | Mean | Difference | p-value |
| **Pre-Stroke mRS** | 40.6590 | 39 | 0.397 | N/A |  |  |  |  |  |
| **Discharge mRS** | 73.3210 | 40 | 0.0002 | N/A |  |  |  |  |  |
| **Initial NIHSS** | 125.5000 | 40 | 9.33E-11 | **46375** (Ref.) | Schererville | 70 | 1.565 |  |  |
|  |  |  |  | **60628** | Chicago | 9 | 18.485 | + 16.92 | 0.0011 |
|  |  |  |  | **46405** | Lake Station | 37 | 8.3917 | + 6.9267 | 0.000155 |
|  |  |  |  | **46324** | Hammond | 110 | 7.4575 | + 5.8925 | 0.0284 |
|  |  |  |  | **46319** | Griffith | 77 | 7.2836 | + 5.7213 | 0.00139 |
|  |  |  |  | **46323** | Hammond | 108 | 6.7572 | + 5.1922 | 0.000103 |
|  |  |  |  | **46312** | East Chicago | 163 | 6.1791 | + 4.6141 | 0.00022 |
|  |  |  |  | **60438** | Lansing | 150 | 5.8149 | + 4.245 | 0.000147 |
|  |  |  |  | **46409** | Gary | 70 | 5.5369 | + 3.9719 | 0.00269 |
|  |  |  |  | **46321** (Ref.) | Munster | 139 | 3.1602 |  |  |
|  |  |  |  | **60628** | Chicago | 9 | 18.485 | + 15.3248 | 0.0103 |
|  |  |  |  | **46405** | Lake Station | 37 | 8.3917 | + 5.2315 | 0.00376 |
|  |  |  |  | **46319** | Griffith | 77 | 7.2836 | + 4.1261 | 0.038 |
|  |  |  |  | **46323** | Hammond | 108 | 6.7572 | + 3.597 | 0.00255 |
|  |  |  |  | **46312** | East Chicago | 163 | 6.1791 | + 3.0189 | 0.0051 |
|  |  |  |  | **60438** | Lansing | 150 | 5.8149 | + 2.6547 | 0.00338 |
|  |  |  |  | **60628** (Ref.) | Chicago | 9 | 18.485 |  |  |
|  |  |  |  | **46304** | Chesterton | 10 | 0.7139 | - 17.771 | 0.0379 |
|  |  |  |  | **46373** | St. John | 27 | 1.9713 | - 16.5137 | 0.0105 |
|  |  |  |  | **46410** | Merrillville | 65 | 3.4777 | - 16.203 | 0.045 |
|  |  |  |  | **46307** | Crown Point | 53 | 4.4328 | - 15.0522 | 0.0171 |
|  |  |  |  | **46405** (Ref.) | Lake Station | 37 | 8.3917 |  |  |
|  |  |  |  | **46373** | St. John | 27 | 1.9713 | - 6.4204 | 0.0369 |
|  |  |  |  | **46307** | Crown Point | 53 | 4.4328 | - 4.9589 | 0.0308 |
| **Discharge NIHSS** |  |  |  |  |  |  |  |  |  |
|  | 75.4920 | 40 | 0.000585 | **46375** (Ref.) | Schererville | 43 | 0.8382 |  |  |
|  |  |  |  | **46312** | East Chicago | 83 | 5.8308 | + 4.9926 | 0.00337 |
|  |  |  |  | **46385** | Valparaiso | 33 | 4.4813 | + 3.6431 | 0.047 |
|  |  |  |  | **46321** (Ref.) | Munster | 67 | 1.2336 |  |  |
|  |  |  |  | **46312** | East Chicago | 83 | 5.8308 | + 4.5972 | 0.0266 |

Based on n=2087 patient data, subset to include all patients who were not transferred from another hospital. Reference (Ref.) ZIP Codes were selected as such to have the least number of total reference groups. Differences among ZIP Code groups were assessed using the Kruskal-Wallis test. Post-hoc pairwise comparisons were performed using Dunn's test (with Bonferroni correction). mRS (modified Rankin Score) data is determined prior to stroke (Pre-Stroke) and upon patient release (Discharge). NIHSS (National Institute of Health Stroke Scale) data is collected upon patient admission (Initial) and release (Discharge).

### II. ZIP Code (Transfers)

Full descriptive statistics of 594 transfer stroke patients are detailed in Supplemental Table S23. Bivariate analysis was not performed. A heatmap of incidence of transfer rates by ZIP Code is displayed in Supplemental Figure S6.

Notably, patients from ZIP Codes with the rates of transfer greater than 5% were ZIP Code 46342, Hobart with 9.06%, ZIP Code 46450, La Porte with 8.51%, ZIP Code 46368, Portage with 8.33%, ZIP Code 46385, Valparaiso with 6.88%, ZIP Code 46360, Michigan City with 6.16%, and ZIP Code 46312, East Chicago with 5.62%.

## DISCUSSION

### I. Result Implications

Focused (25 ZIP Code) Analysis Results: Race and Ethnicity reveal that Black or African American patients presented with higher mean Initial NIHSS and Discharge NIHSS scores than White patients, consistent with previous studies that show higher Initial NIHSS scores in non-Hispanic Black patients compare to non-Hispanic White patients.^26^ These differences may be explained by comorbidities that carry higher 10-year stroke risk such as hypertension and diabetes, which are higher in non-Hispanic Black or African American patients compared to White patients.^27^ A prior study found that Black or African American and Hispanic patients were less likely to be able to identify stroke symptoms compared to White patients.^28^ This difference in health literacy and decreased ability to recognize stroke symptoms may contribute to the higher NIHSS scores upon hospital presentation (discussed further Arrival Mode section).

Focused (25 ZIP Code) Analysis Results: ZIP Code reveals that ZIP Codes with higher mean Initial and Discharge NIHSS scores correlated significantly with ZIP Codes defined by lower median household income, lower education attainment, and higher percentages of minority group populations (i.e., 46312, East Chicago; 46319, Griffith; 46320, 46323, 6324, Hammond; 46342, Hobart; and 46394, Whiting). This is compared to ZIP Codes with higher median household income, higher education attainment, and lower percentages of minority group populations (i.e., 46307, Crown Point; 46321, Munster; and 46375, Schererville). These results are consistent with prior work analyzing the impact of socioeconomic status on stroke severity.^12^ Additionally, it has been found that patients with a lower socioeconomic status and education had a tendency towards worse stroke sign knowledge and Face, Arm, Speech, Time (FAST) public awareness campaign knowledge.^29,30^ The correlation of percentage of minority group populations with stroke scores also validates our earlier findings of the inequities between Race and Ethnicity groups regarding stroke severity. Furthermore, this analysis reveals the unique geographical distribution of these disparities, as seen in the ZIP Code heatmaps. Distance alone, focused within the county borders, does not seem to contribute to stroke severity presentation as significantly as other determinants of health, as seen by the proximity of more severe-scoring ZIP Codes to Stroke Centers, some of them even being locations where the facilities exist (i.e., 46394, Whiting). This disparity relates back to the prior discussion on the impact of healthcare access, health literacy, and other ongoing comorbidities that define stroke recognition and response regardless of distance.

Expanded (46 ZIP Code) Analysis Results: ZIP Code reveals a similar picture to that seen with the 25 ZIP Code analysis, but it reveals that there is an increased stroke severity in non-transfer patients arriving across the state border from Illinois. However, some of the more severe differences are limited by a low count when running the analysis. When looking at the incidence rate distribution, most transfer patients were those who reside in Indiana and who come from farther away ZIP Codes. Notably, patients who reside in ZIP Codes 46342, Hobart and 46312, East Chicago, both contain a Primary Stroke Center and had a high incidence of transfer (greater than 5% across 41 ZIP codes analyzed) compared to other Lake County, IN, ZIP Codes.

Focused (25 ZIP Code) Analysis Results: Payment Source reveals that patients with Medicaid and Medicare had significantly higher scores across all measures compared to patients with Private/HMO/PPO/Other. These results indicate that government-sponsored public insurance programs are associated with increased stroke severity compared to private insurance. This difference in outcome between payment sources was expected and could be explained by unequal healthcare access and treatment. Previous studies have found that patients with private insurance arrived the earliest after symptom onset, had lesser stroke severity by NIHSS at presentation, and had lower in-hospital mortality rates.^31^ Contrary to current literature, Self-Pay/No Insurance had lower mean Discharge mRS scores compared to Medicare – Private/HMO/PPO/Other, although the statistical test had a low power. However, as uninsured patients typically have a lower income, we are unable to make any conclusions from this result without knowing the average socioeconomic status of the Self-Pay/No Insurance group.

Focused (25 ZIP Code) Analysis Results: Mode of Arrival reveals that patients coming from the home/scene who used EMS had significantly higher scores across all four clinical measures compared to patients who took Private transport. This finding demonstrates that patients with more severe strokes at onset opt for EMS utilization over patients with less severe strokes, which aligns with previous findings.^12^ This may be explained by multiple factors, such as the decreased ability to recognize the onset of a stroke when the symptoms at onset are minimal, while severe symptoms at onset may urge EMS usage. Another possibility is that the patient themself or the stroke observer may have the ability to recognize a stroke event but may opt for private transport.^19^ This follows our current understanding that stroke awareness and education in Northwest Indiana are deficient compared to the rest of the United States.^30^ There is a decreasing trend of EMS utilization, which may cause further disparities for patients of lower education. Further awareness of stroke symptoms may increase EMS utilization and produce better stroke outcomes.^19^

### II. Recommendations

#### Individual- and Interpersonal-Level Recommendations

Individuals can make significant improvements to their well-being and minimize their cardiovascular disease risk through lifestyle changes and screenings. Patients are encouraged to discuss with their primary care provider to see what changes make sense for individual health improvement, including blood pressure screening and management, cholesterol and blood sugar control, diet, and exercise. Alongside increased physical activity, patients who seek to stop tobacco use can join smoking cessation classes. Stroke screening assessments, which are offered online or through their healthcare provider, are recommended so that individuals or their loved ones can see their stroke risk and receive informed advice or instructions. Other such interactive digital interventions exist for blood pressure-self management.

#### Organizational-Level Recommendations

Hospitals and health systems can expand data collection and employ SDOH screenings to facilitate further assessment of stroke health and risk factors. Hospitals can also target high-risk patient populations by diversifying their healthcare workforce and providing mobile health clinics to underserved communities. Workplaces can promote employee health by offering regular health screenings and hosting health education workshops. Jointly, physicians are encouraged to speak with their patients regarding their barriers to healthcare access and outline an individualized action plan.

#### Community-Level Recommendations

Stroke awareness programs hosted through hospital stroke education classes or with primary and secondary education centers can inform the public about the symptoms of stroke, what to do when such scenarios occur, and the importance of cardiovascular health. Stroke education campaigns utilizing the FAST (facial droop, arm weakness, speech disturbance, time to call an ambulance) mnemonic has been shown to improve stroke knowledge, encourage prompt action, and ultimately result in improved patient outcomes.^32,33^ In the context of our study population, such an intervention should focus on areas we have previously identified that exhibit lower socioeconomic status and education levels (i.e., 46312, East Chicago; 46319, Griffith; 46320, 46323, 6324, Hammond; 46342, Hobart; and 46394, Whiting).^29^ These communities are also encouraged to increase physical activity by improving public space and transportation accessibility, such as by maintaining sidewalks and bike lanes or by adding point-of-decision prompts to indoor/outdoor spaces to encourage walking and use of stairs.

#### Public Policy-Level Recommendations

Policy initiatives should focus on stroke prevention, early detection, and access to treatment. Expanding insurance coverage for preventative care can reduce the health-related risk factors and comorbidities associated with stroke, especially among low-income patients. Healthy eating can be promoted with government food subsidies for farmers and consumers. Expansion of smoke-free zoning and enforcing smoke-free policies can also limit tobacco usage.

### III. Study limitations

This study was carried out with only two years of patient data, which provides a narrow perspective of patient care and is subject to global trends in health. With 2569 patients for analysis, this study has an appropriate sample size and power when conducting rank-based tests; however, when conducting chi-squared tests and especially any ZIP code analysis, patient counts become low in each cell. While we apply criteria to this data to be appropriate for running the statistical analysis, it must be acknowledged that the sample size could be greater to have a more powerful comparison. The study was further limited due to shortcomings of the retrospective data, including incomplete datasets and the absence of post-discharge mRS information. Certain data points were obtained from national databanks instead of on a by-patient basis, such as Median Household Income and Education Level; thus, certain correlation analyses could not be appropriately controlled to account for confounding variables. Other confounding variables that were not accounted for include EMR-obtainable data such as pre-existing patient co-morbidities or stroke recurrences. The regional specificity of the data also limits the generalizability of the findings of this study to other populations, running the risk of applying a group-level association to individual outcomes.

## CONCLUSIONS

### I. Application of Study Results

Addressing stroke health disparities requires identifying the risk factors associated with more severe clinical outcomes. Through a ZIP Code-level analysis of multiple predictor variables, we were able to better characterize the relationship between social determinants of health and stroke care in Northwest Indiana. Geographic disparities indicate to us that there are regional resource imbalances that can be addressed locally and on a systemic level. Education and income disparities across counties may be addressed through community efforts to make health care more accessible and understandable. This study emphasizes the need to develop targeted interventions that address inequities in the burden of stroke within under-resourced regions.

### II. Future Steps

Our future studies could 1) be updated via an expanded study period and data pool, 2) utilize health outcome data such as 90-day mRS to evaluate stroke care differences, and 3) integrate additional EMR-obtainable data to address confounding variable limitations. Ongoing health descriptors or co-morbidities, such as BMI, hypertension, and diabetes, are more prevalent in minority populations and could be investigated to further delineate the specific stroke risk factors contributing to the disparities in this region. Additionally, clinical measurement of stroke outcome (after treatment) rather than incidence or initial severity may demonstrate more information about differences in care for stroke patients admitted to hospital systems.^34^

## Data Availability

At least one author had full access to all the data in the study and takes responsibility for its integrity and the data analysis.

https://wonder.cdc.gov/mcd.html

https://nccd.cdc.gov/DHDSPAtlas

https://cdc.gov/nchs/nhanes/

## NON-STANDARD ABBREVIATIONS AND ACRONYMS

CSC: Comprehensive Stroke Center
mRS: modified Rankin Scale
NIHSS: National Institutes of Health Stroke Scale
PSC: Primary Stroke Center
ZCTA5: ZIP Code Tabulation Areas

## ACKNOWLEDGEMENTS

We would like to express our gratitude to our project mentor and Urban Medicine Program director Dr. Amy Han, who guided us through this project; to our other leader at Powers Health, Executive Director of Neurosciences Services Dr. Jill Conner, for establishing this ongoing research partnership; and, to the Powers Health Neuroscience Research Data Specialists whose dedicated efforts provided us with the essential data for our project.

## SUPPLEMENTAL MATERIAL

STROBE Reporting Checklist

Disparities Guidelines Checklist

Figures S1-S6

Tables S1-S23

## REFERENCES

1. Tsao CW, Aday AW, Almarzooq ZI, Anderson CAM, Arora P, Avery CL, Baker-Smith CM, Beaton AZ, Boehme AK, Buxton AE, et al. Heart Disease and Stroke Statistics-2023 Update: A Report From the American Heart Association. Circulation. 2023;147:e93–e621. doi: 10.1161/cir.0000000000001123

2. Centers for Disease Control and Prevention and National Center for Health Statistics. Multiple Cause of Death, Provisional Mortality Statistics, CDC WONDER online database. https://wonder.cdc.gov/mcd.html In; 2024.

3. Feigin VL, Stark BA, Johnson CO, Roth GA, Bisignano C, Abady GG, Abbasifard M, Abbasi-Kangevari M, Abd-Allah F, Abedi V, et al. Global, regional, and national burden of stroke and its risk factors, 1990&#x2013;2019: a systematic analysis for the Global Burden of Disease Study 2019. The Lancet Neurology. 2021;20:795–820. doi: 10.1016/S1474-4422(21)00252-0

4. Stierman B, Afful J, Carroll MD, Chen T-C, Davy O, Fink S, Fryar CD, Gu Q, Hales CM, Hughes JP, et al. National Health and Nutrition Examination Survey 2017–March 2020 Prepandemic Data Files Development of Files and Prevalence Estimates for Selected Health Outcomes. In: Series : NCHS National Health Statistics Reports. Hyattsville, MD: 10.15620/cdc:106273; 2021.

5. Centers for Disease Control and Prevention and National Center for Health Statistics. National Health and Nutrition Examination Survey (NHANES) public use data files. www.cdc.gov/nchs/nhanes/. 2024. Accessed June 14.

6. Indiana State Department of Health [database online]. Indianapolis, IN: ’Updated’ June 14, 2020.

7. Centers for Disease Control and Prevention. Atlas of Heart Disease and Stroke. https://nccd.cdc.gov/DHDSPAtlas. 2021. Accessed March 27.

8. Indiana Department of Health Tobacco Prevention and Cessation. State Fiscal Year 2023 Report. In; 2023.

9. Centers for Disease Control and Prevention Population Health Surveillance Branch [database online]. 2021.

10. Marmot M, Friel S, Bell R, Houweling TAJ, Taylor S. Closing the gap in a generation: health equity through action on the social determinants of health. The Lancet. 2008;372:1661–1669. doi: 10.1016/S0140-6736(08)61690-6

11. Havranek EP, Mujahid MS, Barr DA, Blair IV, Cohen MS, Cruz-Flores S, Davey-Smith G, Dennison-Himmelfarb CR, Lauer MS, Lockwood DW, et al. Social Determinants of Risk and Outcomes for Cardiovascular Disease. Circulation. 2015;132:873–898. doi: doi:10.1161/CIR.0000000000000228

12. Yadav RS, Chaudhary D, Avula V, Shahjouei S, Azarpazhooh MR, Abedi V, Li J, Zand R. Social Determinants of Stroke Hospitalization and Mortality in United States’ Counties. J Clin Med. 2022;11. doi: 10.3390/jcm11144101

13. U.S. Census Bureau. American Community Survey 5-Year Estimates (ZCTA5). In; 2022.

14. U.S. Census Bureau. Decennial Census. In; 2020.

15. The Joint Commission. Find Accredited Organizations. https://www.jointcommission.org/who-we-are/who-we-work-with/find-accredited-organizations/. 2025. Accessed March.

16. Banks JL, Marotta CA. Outcomes validity and reliability of the modified Rankin scale: implications for stroke clinical trials: a literature review and synthesis. Stroke. 2007;38:1091–1096. doi: 10.1161/01.STR.0000258355.23810.c6

17. Kazi SA, Siddiqui M, Majid S. Stroke Outcome Prediction Using Admission Nihss In Anterior And Posterior Circulation Stroke. J Ayub Med Coll Abbottabad. 2021;33:274–278.

18. Adams HP, Jr., Davis PH, Leira EC, Chang KC, Bendixen BH, Clarke WR, Woolson RF, Hansen MD. Baseline NIH Stroke Scale score strongly predicts outcome after stroke: A report of the Trial of Org 10172 in Acute Stroke Treatment (TOAST). Neurology. 1999;53:126–131. doi: 10.1212/wnl.53.1.126

19. Astasio-Picado Á, Chueca YC, López-Sánchez M, Lozano RR, González-Chapado MT, Ortega-Trancón V. Analysis of the Factors Intervening in the Prehospital Time in a Stroke Code. J Pers Med. 2023;13. doi: 10.3390/jpm13101519

20. Ormseth CH, Sheth KN, Saver JL, Fonarow GC, Schwamm LH. The American Heart Association’s Get With the Guidelines (GWTG)-Stroke development and impact on stroke care. Stroke Vasc Neurol. 2017;2:94–105. doi: 10.1136/svn-2017-000092

21. von Elm E, Altman DG, Egger M, Pocock SJ, Gøtzsche PC, Vandenbroucke JP. The Strengthening the Reporting of Observational Studies in Epidemiology (STROBE) Statement: Guidelines for reporting observational studies. International Journal of Surgery. 2014;12:1495–1499. doi: 10.1016/j.ijsu.2014.07.013

22. Aoki J, Suzuki K, Kanamaru T, Kutsuna A, Katano T, Takayama Y, Nishi Y, Takeshi Y, Nakagami T, Numao S, et al. Association between initial NIHSS score and recanalization rate after endovascular thrombectomy. J Neurol Sci. 2019;403:127–132. doi: 10.1016/j.jns.2019.06.033

23. Esri. USA Zip Code Boundaries. In; 2024.

24. Ahmad OB, Boschi Pinto C, Lopez AD. Age Standardization of Rates: A New WHO Standard. GPE Discussion Paper Series*: No* 31. 2001:10–12.

25. R Core Team. R: A Language and Environment for Statistical Computing; Vienna, Austria: R Foundation for Statistical Computing; 2024.

26. Jones EM, Okpala M, Zhang X, Parsha K, Keser Z, Kim CY, Wang A, Okpala N, Jagolino A, Savitz SI, et al. Racial disparities in post-stroke functional outcomes in young patients with ischemic stroke. J Stroke Cerebrovasc Dis. 2020;29:104987. doi: 10.1016/j.jstrokecerebrovasdis.2020.104987

27. Cushman M, Cantrell RA, McClure LA, Howard G, Prineas RJ, Moy CS, Temple EM, Howard VJ. Estimated 10-year stroke risk by region and race in the United States: geographic and racial differences in stroke risk. Ann Neurol. 2008;64:507–513. doi: 10.1002/ana.21493

28. Ellis C, Egede LE. Ethnic disparities in stroke recognition in individuals with prior stroke. Public Health Rep. 2008;123:514–522. doi: 10.1177/003335490812300413

29. Rioux B, Brissette V, Marin FF, Lindsay P, Keezer MR, Poppe AY. The Impact of Stroke Public Awareness Campaigns Differs Between Sociodemographic Groups. Can J Neurol Sci. 2022;49:231–238. doi: 10.1017/cjn.2021.76

30. Raab C. The Use of a Stroke Specific Community Needs Assessment Survey to Drive Stroke Education in Underserved Communities (4905). Neurology. 2021;96:4905. doi: doi:10.1212/WNL.96.15_supplement.4905

31. Medford-Davis LN, Fonarow GC, Bhatt DL, Xu H, Smith EE, Suter R, Peterson ED, Xian Y, Matsouaka RA, Schwamm LH. Impact of Insurance Status on Outcomes and Use of Rehabilitation Services in Acute Ischemic Stroke: Findings From Get With The Guidelines-Stroke. Journal of the American Heart Association. 2016;5:e004282. doi: doi:10.1161/JAHA.116.004282

32. Amano T, Yokota C, Sakamoto Y, Shigehatake Y, Inoue Y, Ishigami A, Hagihara T, Tomii Y, Miyashita F, Toyoda K, et al. Stroke education program of act FAST for junior high school students and their parents. J Stroke Cerebrovasc Dis. 2014;23:1040–1045. doi: 10.1016/j.jstrokecerebrovasdis.2013.08.021

33. American Heart Assocation. Stroke Symptoms. https://www.stroke.org/en/about-stroke/stroke-symptoms. 2025.

34. van Swieten JC, Koudstaal PJ, Visser MC, Schouten HJ, van Gijn J. Interobserver agreement for the assessment of handicap in stroke patients. Stroke. 1988;19:604–607. doi: 10.1161/01.str.19.5.604

